# Predicted Infection Risk for Aerosol Transmission of SARS-CoV-2

**DOI:** 10.1101/2020.10.08.20209106

**Authors:** Martin Kriegel, Udo Buchholz, Petra Gastmeier, Peter Bischoff, Inas Abdelgawad, Anne Hartmann

## Abstract

Currently, the respiratory route is seen as the most important transmission path for SARS-CoV-2. In this investigation, models of other researchers which had the aim of predicting an infection risk for exposed persons in a room through aerosols emitted by an infectious case-patient were extended. As a novelty – usually neglected – parameters or boundary conditions, namely the non-stationarity of aerosols and the half-life of the aerosolized virus, were included and a new method for determining the quanta emission rate based on measurements of the particle emission rate and respiratory rate at different types of activities was implemented.

As a second step, the model was applied to twelve outbreaks to compare the predicted infection risk with the observed attack rate. To estimate a “credible interval” of the predicted infection risk, the quanta emission rate, the respiratory rate as well as the air volume flow were varied.

In nine out of twelve outbreaks, the calculated predicted infection risk via aerosols was found to be in the range of the attack rate (with the variation of the boundary conditions) and reasons for the observed larger divergence were discussed.

The validation was considered successful and therefore the use of the model could be recommended to predict the risk of an infection via aerosols in given situations. Furthermore, appropriate preventive measures can be designed.

## Introduction

The respiratory route is the main mode of transmission for the virus causing COVID-19 (SARS-CoV-2) [1, 2, 3]. The virus is transported on particles that can enter the respiratory tract. Whereas larger particles (droplets) are only able to stay in the air for a short time and just in the near field (approx. 1.5 m), due to rapid settling, smaller particles (called aerosols) are also concentrated in the near field and in addition can follow the air flow and cause infections in the far field. Epidemiologically, short-range transmission (through aerosols or droplets) is distinguished from long-range transmission (aerosol).

In order to perform an infection risk assessment for the airborne transmission in the far field and to introduce appropriate preventive measures, it would be necessary to know the amount of aerosols produced by an infected person during various activities, how many viruses stick to the aerosols and how many viruses are necessary to cause an infection. However, this information is usually available only very late in the course of a pandemic, if it can be determined at all. Another well-known approach is to use retrospective analysis of infection outbreaks that are very probably due to far field transmission to determine a virus-laden aerosol concentration. Exposed people have inhaled the virus-laden aerosols according to their respiratory volume flow. The approach presented here corresponds to a combination of known measured source rates of respiratory aerosols at different activities and the retrospective analysis of previous infection incidents with the aim to calculate a predicted infection risk via aerosols and to calculate the necessary air volume flow to reduce the risk of COVID-19 infections.

## Current state

The so-called aerosols (liquid or solid particles in a dispersed phase with a fluid) as well as droplets differ by size. The particles, which are transported in a fluid over a longer distance, are called aerosols. Droplets are more strongly influenced by gravitation and are deposited more rapidly. The size of particles which can be transported in air for a longer distance varies with the velocity of the fluid. In internal spaces with typical air velocities of up to 0.2 m/s particles smaller than 10 μm will be distributed by air very well; with a higher air velocity, larger particles may also be transported in air.

SARS-CoV-2 was found to be transmitted via close contact as well as over distance in internal spaces, whereas in distant transmission so-called super-spreading events are more probable [1, 2, 4].

In 1978, Riley et al. [5] evaluated a measles outbreak in a suburban elementary school. Based on the number of susceptible persons (S), which have been infected (D) during each stage of infection, the risk (P) for an infection in this stage has been calculated with equation (1). Therefore, the risk for an infection has been defined as the percentage of infected persons from the number of pupils not already infected or vaccinated.

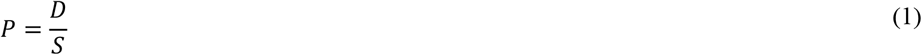

A Poisson-distribution of the risk of infection has been assumed as well as a stationary and evenly distributed concentration of the pathogens in the room air. Equation (2) shows the Poisson-distribution.

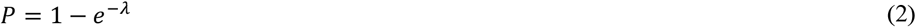

Therefore, Wells defined in 1955 [6] a size called quantum as the number of emitted infectious units, where the probability to get infected is 1 − *e*^−1^ = 63.2%. Hence, a quantum can be seen as a combination of the number of emitted aerosols with the virus transported on them and a critical dose which may result in an infection in 63.2 % of the exposed persons. Riley [5] combined the quantum concept with equation (2) to produce equation (3).

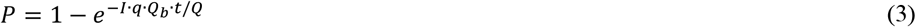

In equation (3), the number of infectious persons (I), the quanta emission rate depending on the activity (q), the pulmonary ventilation rate of exposed susceptible persons (Q_b_), the duration of stay (t) and the volume flow of pathogen free air (Q) was used. The quotient q/Q represents the quanta concentration.

In poorly ventilated rooms, the assumption of a stationary concentration of quanta is not justified, because of the amount of time necessary before a stationary concentration is reached. The normalized time-dependent concentration process can be calculated according to equation (4) and is shown in Figure 1. How rapidly the concentration of a human emitted contamination in a room raises depends on the air exchange rate (ACH) and the time (t). This relative concentration (c_rel_) can be seen as an increase in the concentration compared to the volume flow.

**Figure 1:**
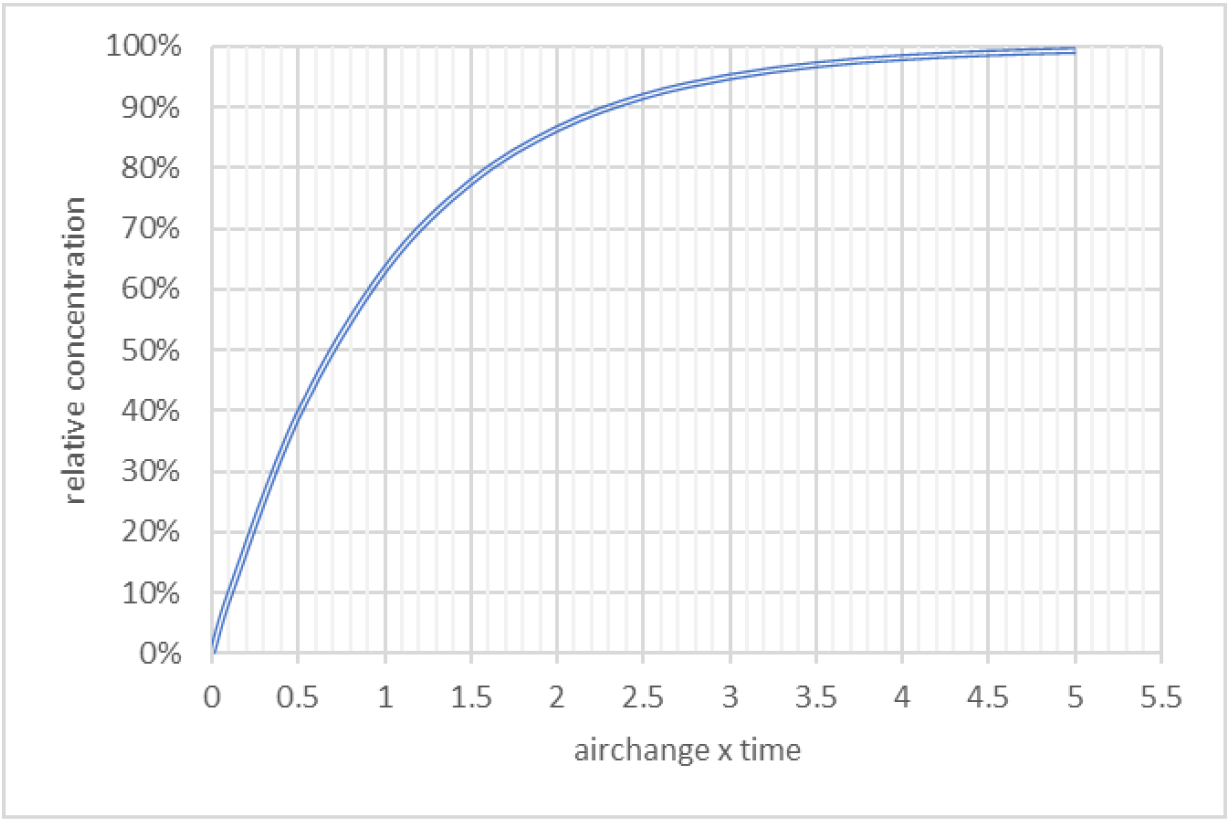
Relative concentration curve as a function of air exchange rate and time.

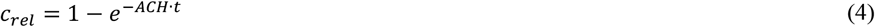

In all published studies identified, ideal mixing ventilation was assumed, which means that aerosols are evenly distributed in the room air. To avoid this assumption Noakes and Sleigh [7] divided the room air into different zones, which are themselves considered to be well mixed and have a uniform concentration. This should make it possible to calculate local differences in concentration and thus locally differing infection risks. Furthermore, other studies, which focus on the unsteady conditions mostly use the boundary condition of a starting concentration of 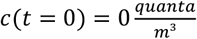. Gammaitoni and Nucci [8] implemented the starting condition of *c*(*t* = 0) = *c*_0_ as well as the number of exposed susceptible people, which may also change over time depending on their immune status.

To estimate the risk of infection in a given setting by a given infectious person with the Wells-Riley-equation either the quanta emission rate or the P have to be known. In the beginning of an epidemic, both values are unknown. Dai and Zhao [9] correlated and calculated q for SARS-CoV-2 based on the basic reproduction number (R_0_) known from former outbreaks of MERS, tuberculosis, influenza and SARS-CoV-1 and published the equation (5). If R_0_ is known, q can be estimated as proposed by Dai and Zhao [9]. For SARS-CoV-2 the average basic reproduction number has been estimated to be 3.28 [10], 3.32 [11] and 3.77 [12].

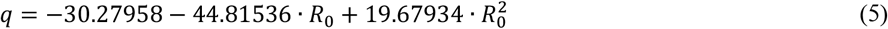

In various studies of infection occurrences associated with SARS-CoV-2, q was determined using the Wells-Riley equation. Different authors [9, 13] found a range of 22 to 61 quanta/h with an assumed low activity (breathing, speaking) and values of 341 to 1190 quanta/h when singing.

The virus can be transported on particles in air and the emission of aerosols can be used as an indicator for the emission of the virus, but a correlation between q and the aerosol emission rate (E) has not been investigated so far. In measurements at the Hermann-Rietschel-Institute (HRI) of Technical University of Berlin [14, 15] the particle emission rates during breathing, speaking, coughing as well as singing was measured. During breathing through the nose about 25 particles/s was emitted and during coughing about 13,700 particles/cough. It is therefore evident that depending on the activity a wide range of particle emission rates can be found. The transmission of a pathogen via aerosols is also influenced by the stability of the virus in the environment. In an experimental study van Doremalen et al [16] measured the decrease of infectious virus in the air and on different surfaces and compared SARS-CoV-1 and SARS-CoV-2. Under the given conditions, the half-life of SARS-CoV-2 as well as SARS-CoV-1 was about 1.1 h.

The longer aerosols stay in the room air, the more the proportion of inactivated virus increases. The age of air is dependent on the airflow as well as the position of the source of the pollutants. At each point in the room, a local age of air can be calculated. The age of air (τ_n_) can be used as a measure to evaluate the air quality. For an ideal mixing ventilation, the mean age of air is equal to the nominal time constant, which can be calculated using equation (6) as the quotient of the room volume (V) and the air volume flow (Q).

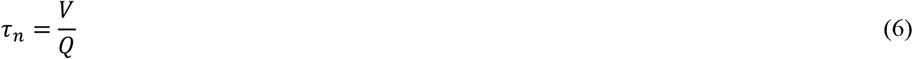

Besides the number of emitted pathogen-laden aerosols, the number of inhaled pathogens also plays an important role, with regard to the assessment of the risk of infection. The pulmonary ventilation rate may differ with different activities. Gupta et al. [17] performed a study with 25 healthy adults and found that the volume flow fluctuated in the form of a sine wave during simple breathing, but gave a more constant volume flow during talking.

In measurements with athletes as well as sedentary persons a maximum volume flow for the athletes of 200 l/min (12 m^3^/h) was found by Córdova and Latasa [18].

To measure the airflow without movement restrictions, a helmet was used by Jiang et al. [19] in 32 subjects (16 males, 16 females) during speaking with different volumes as well as during singing.

A comparison between a machine-learning based model and measurements of respiratory rate was performed by Dumond et al. [20].

As a conclusion, the following average values can be used for adults:

- low activity (breathing while lying): 0.45 m^3^/h [19]
- low activity (breathing while sitting, standing or talking): 0.54 m^3^/h [19, 20]
- singing: 0.65 m^3^/h [21]
- mid activity (physical work): 0.9 m^3^/h [20]
- sports: 1.2 m^3^/h [18, 20]

For children, the lung volume is smaller. Therefore, the respiratory rate for children aged 14 can be assumed to be 0.45 m^3^/h for low activity (breathing while sitting, standing, talking) [22].

## Methods

### Extension of the Wells-Riley equation for calculating the Predicted Infection Risk via Aerosols (PIRA)

The Wells-Riley equation can be summarized as equation (7). To calculate the predicted infection risk via aerosols (PIRA) in the far field of a room the concentration of quanta (c(t)) and the respiratory rate (Q_b_) must be known. The integration of c(t) can be understood as the number of particles inhaled per m^3^/h. Together with Q_b_, the number of inhaled quanta can therefore be calculated.

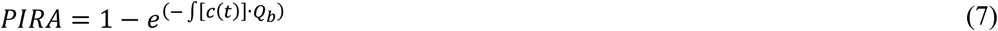

Equation (5) can be used for the definition of the quantum emission. This leads to a quanta emission rate of q = 40 1/h at an assumed mean R_0_ = 3.35. The mathematical approximation presented by Dai und Zhao [9] can be optimized by equation (8), see Figure 2. For Figure 2 the quanta emission rate has been correlated with R_0_ of tuberculosis [23, 24], Influenza [23, 25], MERS [26, 27] and SARS-CoV [23, 28].

**Figure 2:**
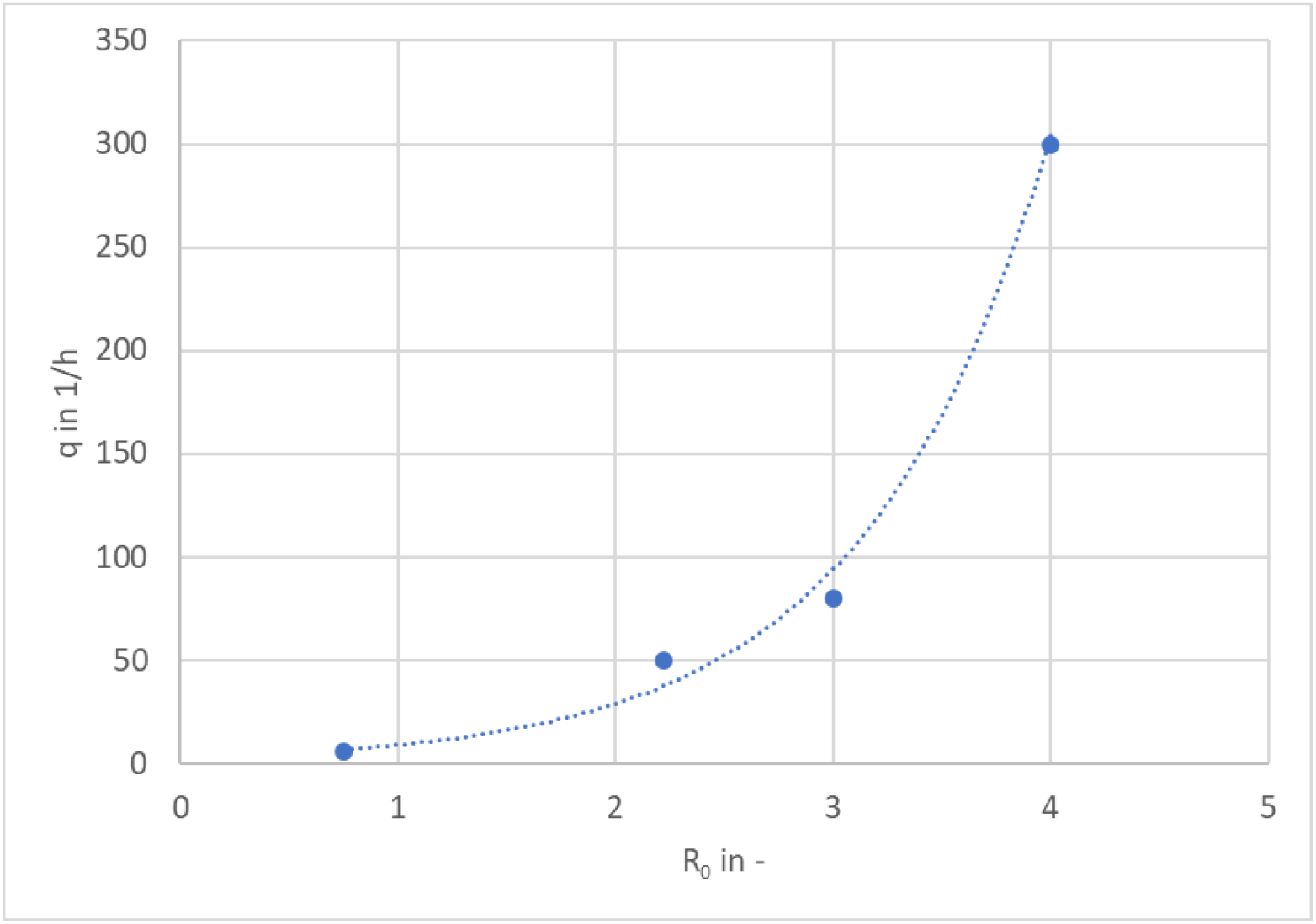
Relationship between quanta emission rate and R_0_ according Dai and Zhao [9].

Equation (8) results in q = 139 1/h for an assumed mean R_0_ = 3.35. Because of the high variance of the q as well as R_0_ given in the currently available literature the difference between the q calculated regarding equation (5) and equation (8) seems reasonable.

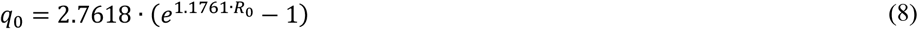

q is influenced by the activity of the person as was shown by Buonanno et al. [29]. Therefore, the measured aerosol emission rates *E* [14, 15] were correlated with the calculated quanta emission rates influenced by the activity *q*_*a*_ by equation (9). For low-activity (breathing, talking, sitting, standing) a basic volume flow Q_b,o_ and normal activity = low activity (breathing, talking, sitting, standing) with a basic emission rate of E_0_ was used. Furthermore, the basic q (q_0_) was calculated with usage of R_0_ regarding equation (8). With these specifications q_a_ can be calculated.

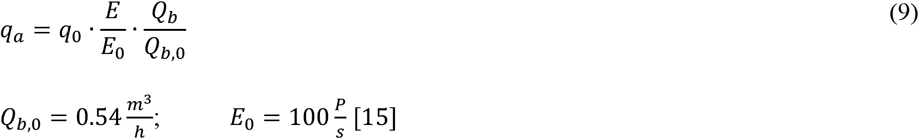

The effect of e.g. mouth-nose protection can be considered by using their filtration efficiency (F_MNS_) like in equation (10) which however will not be further considered in the following.

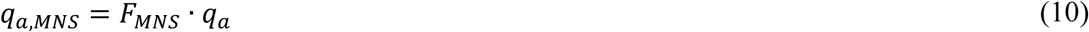

It is known that the infectivity of an infected person depends on the disease progression over time [30]. This is shown qualitatively in Figure 3. With a simplified mathematical approach, this can be integrated into the quanta source rate. An equation could be implemented to take this into account.

**Figure 3:**
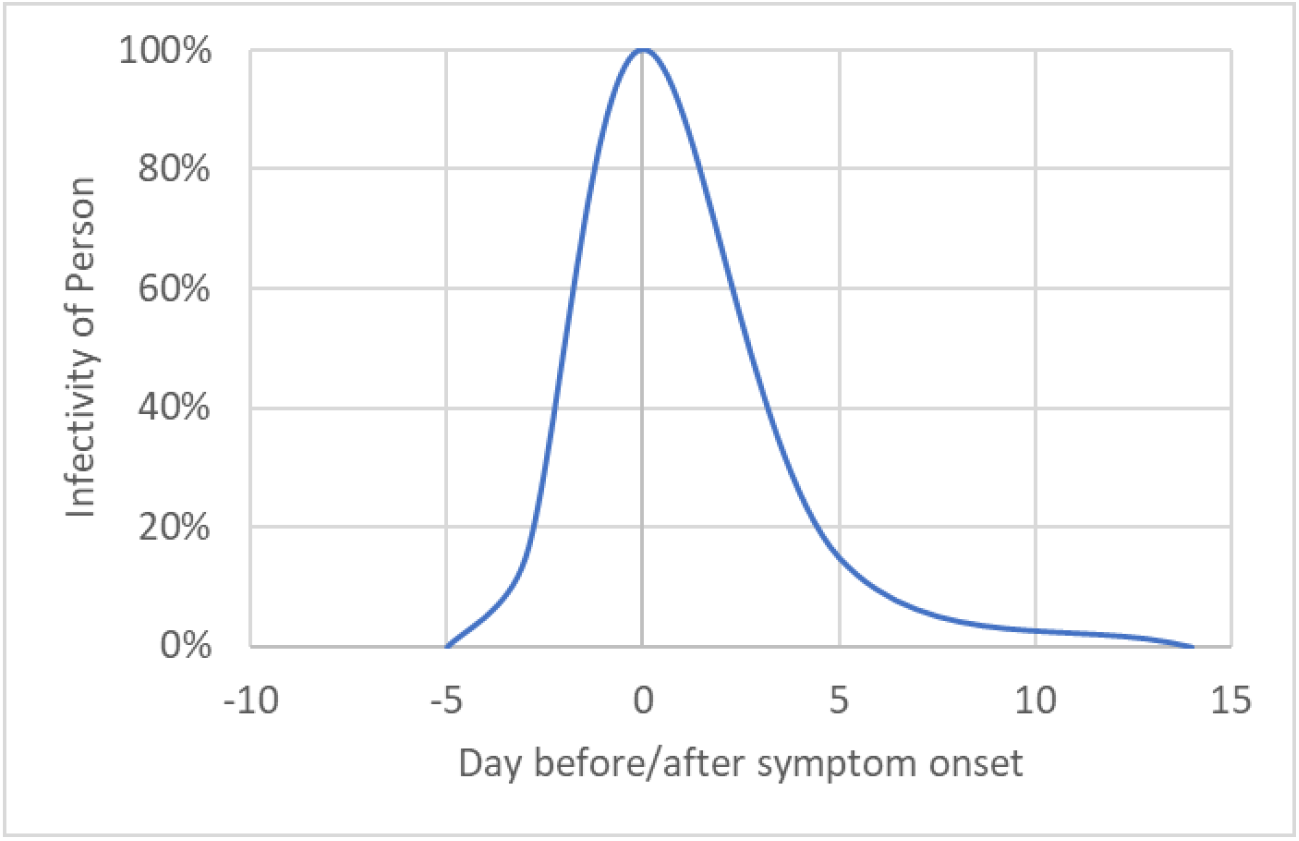
Infectivity depending on the disease progression.

Under experimental conditions, the half-life of virus was measured as 1.1 h [16]. Thus, the number of emitted infectious quanta at time t (q_a_(t)) is calculated according to equation (11).

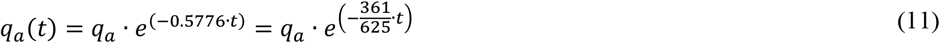

The concentration of quanta during the increase c_I_(t) can be calculated according to equation (12) with the number of infectious persons (n).

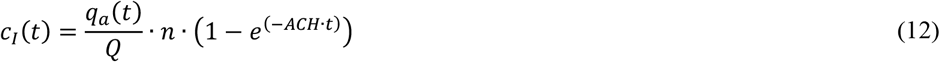

An additional case is considered that if the time t is longer than the age of the air τ_n_, most of the virus-laden aerosols have left the room with the exhaust before the inactivation can take place. Therefore, this concentration during the steady state situation is called c_τ_(t).

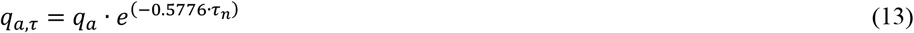

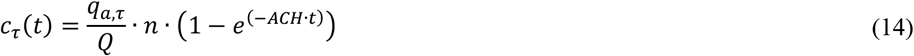

with τ_n_ regarding equation (6)

For calculating the risk of infection the unsteady concentrations c_I_ and c_τ_ has to be used, to include the time-dependent increase in concentration and to include the time-dependent viability of the virus, otherwise the result could be overestimated or underestimated.

For equation (7) the integration of c_I_(t) and c_τ_(t) is necessary.

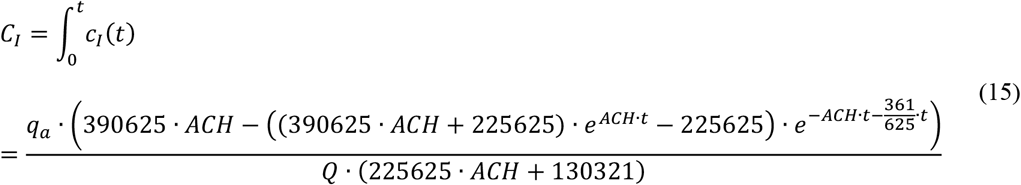

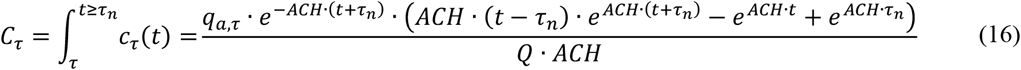

The Predicted Infection Risk via Aerosols can be calculated by equation (17) and (18).

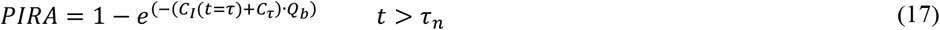

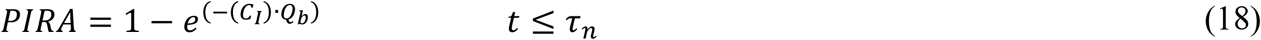

For the calculation of PIRA the following assumptions must be considered:

- the aerosols are ideally mixed in the room
- the near field (up to approx. 1.5 m distance from the emitting person) can contain a much higher virus-laden aerosol concentration
- the air, which is introduced into the room, is free of virus-laden aerosols (e.g. outside air)
- no deposition of small particles is considered, because the settling time is longer than the stability of the virus and the deposition rate would therefore be substantially smaller than the inactivation
- the concentration of aerosols at the beginning is 0

## Results

The PIRA calculation model was validated by using parameters of several known outbreaks during the SARS-CoV-2 pandemic. Twelve different scenarios either scientifically published or registered by the local health authorities were selected (A-L). The boundary conditions for the calculations of these situations can be found in Table 1.

**Table 1:**
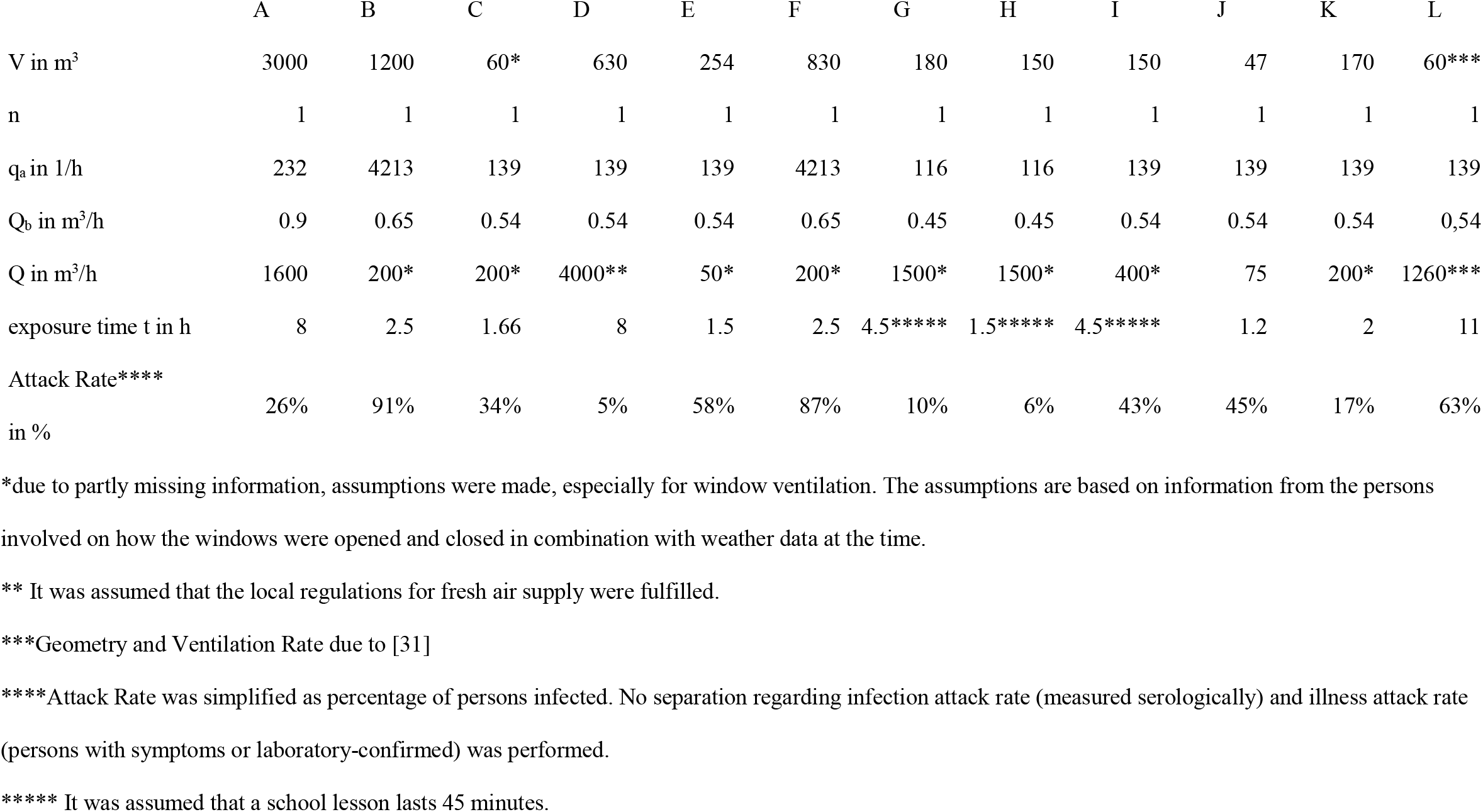
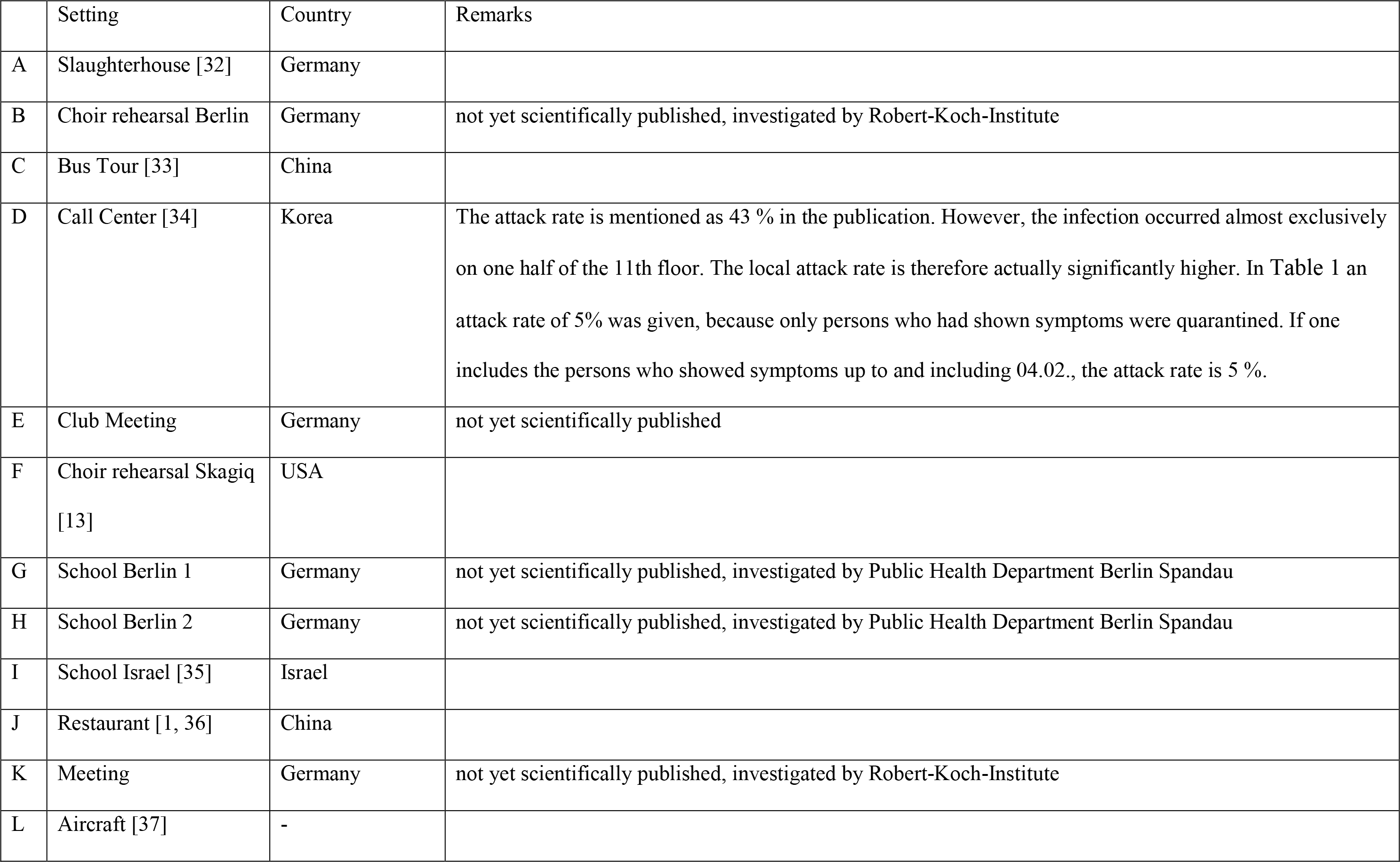
Boundary conditions of SARS-CoV-2 outbreaks for the retrospective calculation of PIRA.

The infection events used for the validation of the model are shown in Table 1 with the necessary parameters for the calculation. In the following, the comparison between the documented Attack Rate (AR) and the PIRA is drawn.

The q used here was calculated according to equation (9) with the assumption that the cases emitted particles as measured in [14, 15]. Due to the high spread of the particle emission E and the unknown proportions of breathing, speaking, singing and shouting as well as the respiratory volume flows, simplified a-priori assumptions were made. To take into account the effects of the uncertainties regarding q, Q_b_ and especially with window ventilation on Q, these values were further varied - q by +/- 20%, Q_b_ by +/- 20% and Q by +/- 50%, individually and in combination, which then lead to a minimum PIRA and maximum PIRA. Figure 4 presents PIRA and the minima and maxima calculated with the different variants. The red dots show the documented AR.

**Figure 4:**
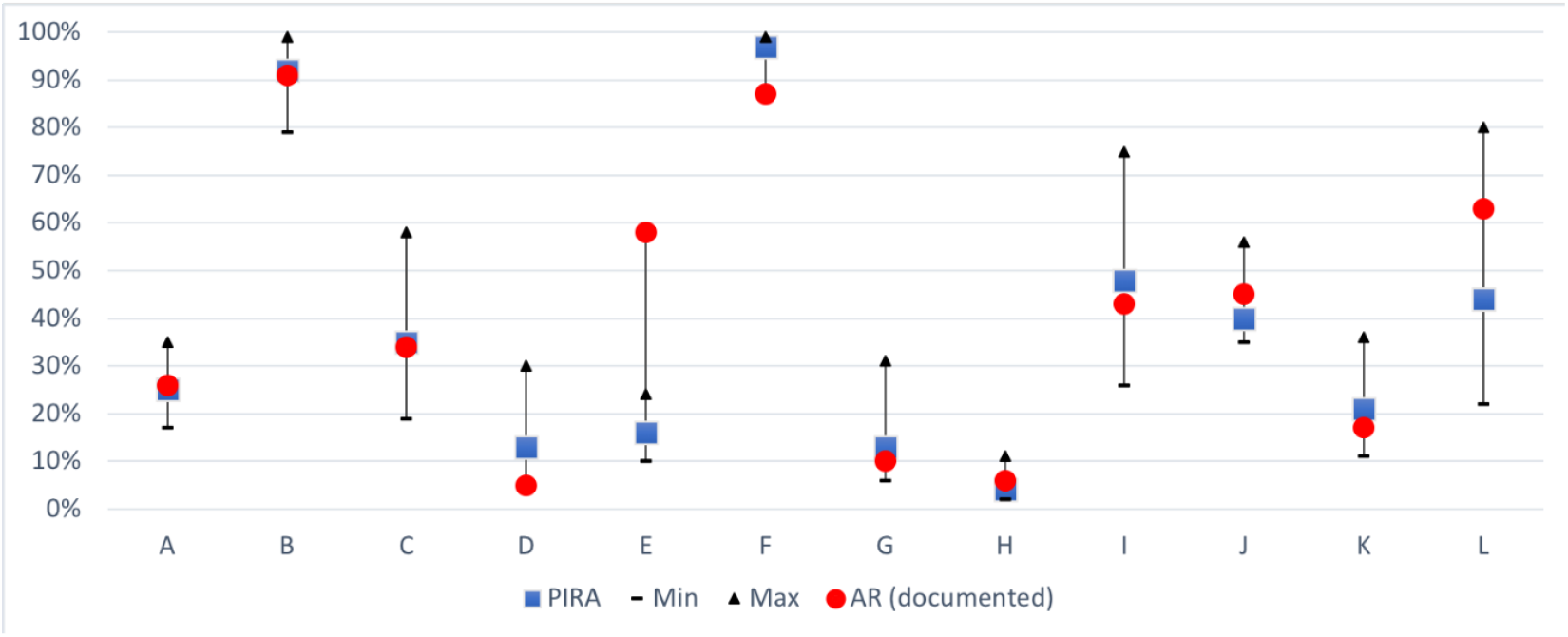
Comparison between PIRA with the variants Min and Max to the documented AR assuming that all cases were caused by long-range transmission

In nine out of twelve outbreaks, the attack rate lies in the Min Max values of the calculated PIRA (see Table 2).

**Table 2:** Results of the Calculation of PIRA and comparsion with the documented AR.

|  | A | B | C | D | E | F | G | H | I | J | K | L |
| --- | --- | --- | --- | --- | --- | --- | --- | --- | --- | --- | --- | --- |
| PIRA | 25% | 92% | 35% | 13% | 16% | 97% | 13% | 4% | 48% | 40% | 21% | 44% |
| Min | 17% | 79% | 19% | 6% | 10% | 88% | 6% | 2% | 26% | 35% | 11% | 22% |
| Max | 35% | 99% | 58% | 30% | 24% | 99% | 31% | 11% | 75% | 56% | 36% | 80% |
| AR (documented) | 26% | 91% | 34% | 5% | 58% | 87% | 10% | 6% | 43% | 45% | 17% | 63% |

From the PIRA model, it can be calculated how much volume flow per hour of exposure time is required to not exceed a certain PIRA. The results are shown in Figure 5. It can be seen that for a PIRA of 10% a volume flow of clean air of 750 m^3^/h and hour of exposure has to be supplied to the room (see Table 3), whereas for two hours 1500 m^3^/h will be necessary for the same PIRA.

**Table 3:** Required volume flow at a certain exposure time for a defined PIRA

|  |  | PIRA |  |  |
| --- | --- | --- | --- | --- |
|  |  | 1% | 5% | 10% |
| exposure<br>time in h | 1 | 7500 m³/h | 1500 m³/h | 750 m³/h |
|  | 2 | 15000 m³/h | 3000 m³/h | 1500 m³/h |
|  | 3 | 22500 m³/h | 4500 m³/h | 2250 m³/h |

**Figure 5:**
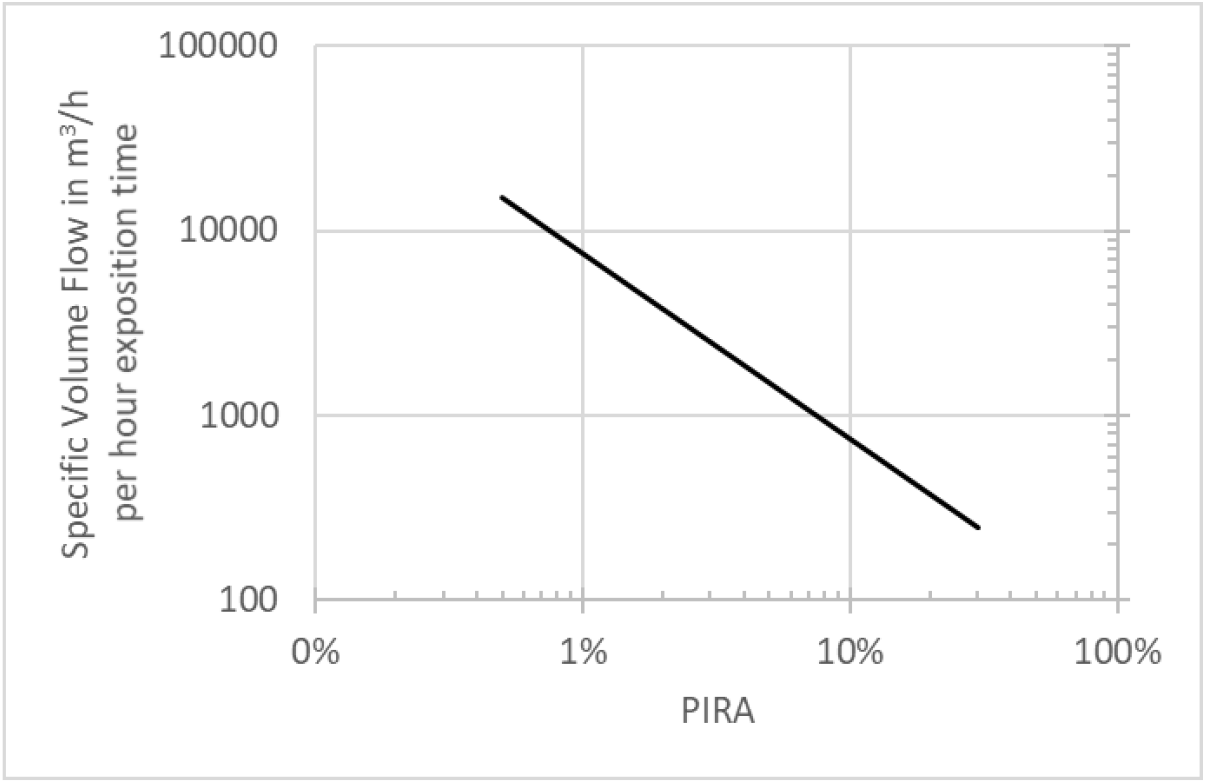
Required specific volume flow per hour exposure time with regard to meeting a specific PIRA

As a regression of the calculated results presented in Figure 5, equation (19) was derived. Using equation (19) the required volume flow per hour of exposure time can be calculated. This information refers to the steady state if the product of ACH and t is higher than 5.0, see Figure 1. If the product is smaller, correspondingly lower volume flows lead to the respective PIRA.

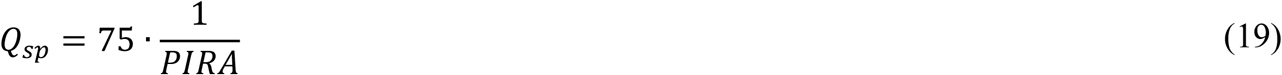

Table 3 lists practical examples of the required volume flows depending on the exposure time and PIRA.

Another type of evaluation shows the possible number of infected persons in relation to the person-related volume flow and the number of persons in a room (see Figure 6), if several exposed persons are in a room and each person would have 6 m^3^ of room volume available. If more volume flow is available to each person, the result changes marginally and tends towards a lower PIRA. For an exposure time of two hours a volume flow of 20 m^3^ /(h·Per) resulted in five probably infected persons in a room with 20, 40 or 100 persons, but of course only one probably infected person in a room with two persons. For a volume flow of 60 m^3^/(h·Per) the number decreases to two probably infected persons in a room with 20, 40 or 100 persons, but of course stayed at one probable person for just two persons in a room.

**Figure 6:**
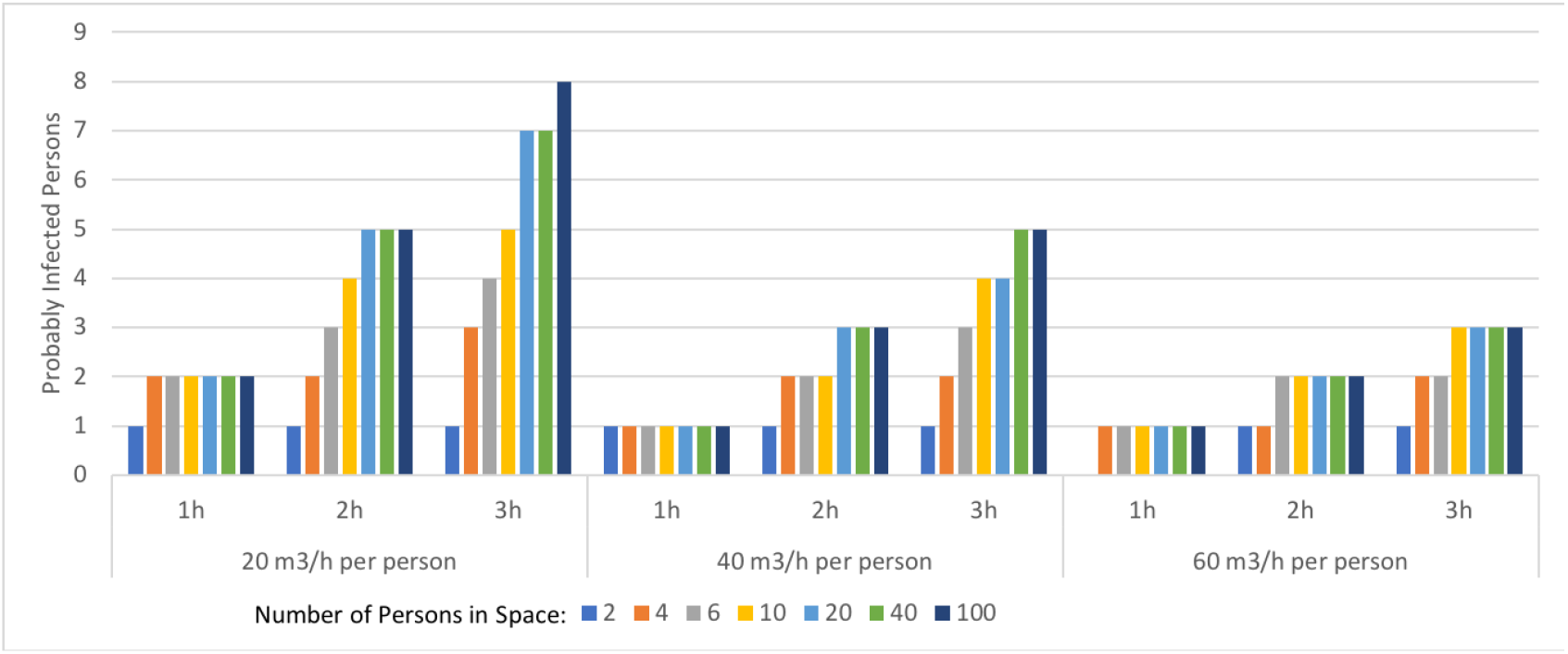
Number of persons probably infected according to a specific volume flow.

## Discussion

For the first time with a q based on R_0_ the PIRA was calculated for different known SARS-CoV-2 outbreaks. In addition, the necessary air volume flows to reduce the risk of infection were calculated.

In Table 4 the q-values are compared according to the stationary equation (3) and the PIRA model under the boundary conditions of Table 1. Large differences between the same activities were observed in the calculation of the steady-state.

**Table 4:**
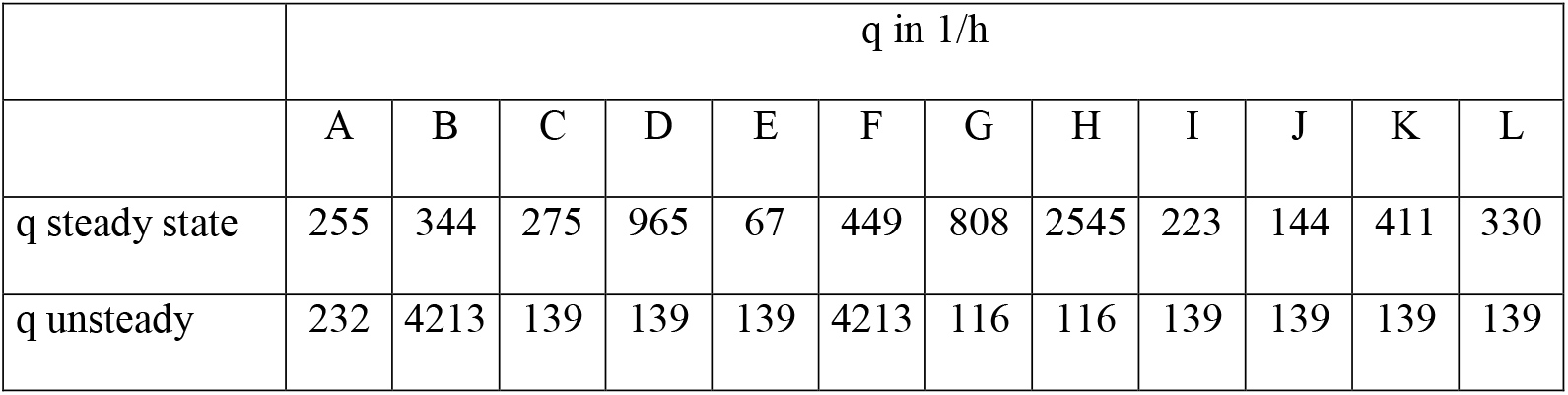
comparison steady state and unsteady calculated quanta.

|  | q in 1/h |  |  |  |  |  |  |  |  |  |  |  |
| --- | --- | --- | --- | --- | --- | --- | --- | --- | --- | --- | --- | --- |
|  | A | B | C | D | E | F | G | H | I | J | K | L |
| q steady state | 255 | 344 | 275 | 965 | 67 | 449 | 808 | 2545 | 223 | 144 | 411 | 330 |
| q unsteady | 232 | 4213 | 139 | 139 | 139 | 4213 | 116 | 116 | 139 | 139 | 139 | 139 |

First of all, in comparison, with fixed q according to equation (9) very good agreement can be achieved with the documented AR in the retrospectively considered cases A, B, C, H, I, J, K.

In outbreak D, the documentation does not clearly show how large the actual AR was for the area under consideration. Due to the fact, that only the person who showed symptoms was isolated, it is possible that in the meantime, additional persons, who were already infected by the index case may have infected others (e.g. pre-symptomatically) leading to the high AR. In outbreak E, there is little documentation of the infection process, and further contact between some of the persons had occurred in a restaurant afterwards. Furthermore, it has not been determined whether the infection can be attributed to only one person. The high AR after a short time of exposure allows the conclusion that either two index persons were present or that the exposure time was prolonged by the meeting in a restaurant.

In outbreak F, the group was not together for the entire time and some of the subjects continued rehearsing in another room. For this reason, the exposure time for the whole group was lower and this may account for the lower AR than calculated by PIRA.

In outbreak L an air exchange rate of 21 1/h was assumed. A relatively small change in the assumed volume flow has a significant influence on the result of PIRA (where the total exposure time was used). Furthermore, it cannot be excluded that droplet transmission may also have happened.

Secondly, many assumptions were made, therefore it is not clear if the formula is already optimal, perhaps further optimization during the course of the pandemic is possible if further knowledge becomes available.

Third, the calculation model does not consider the sedimentation behavior of particles. It is known that at higher air velocities, and especially at high turbulence, the sedimentation behavior increases. In typical indoor air flows this decrease is about 10 % per hour. Compared to the uncertainty of the overall emission rate, this effect is not significant.

Fourth, the calculation model assumes a homogeneous distribution of the particles in the room air. Practically however, the ventilation effectiveness is locally very different. The differences can be slightly greater than 100%.

Finally, it must be noted that the aerosol concentration is significantly higher in the near field of the emitting person and the results of PIRA are not valid within the generally accepted 1.5 m distance rules.

## Conclusion

It was shown in this investigation that it was possible to calculate the risk of an infection via aerosols for situations where the long-distance transmission is more important. By using the model presented here, a good agreement to previous infection outbreaks in different settings and different attack rates was achieved. Previous retrospectively determined quanta emission rates usually assumed a stationary state. However, if the concentration process is important for the total amount of inhaled virus-laden aerosols (usually at ACH x t < 5), then a stationary observation leads to an incorrect boundary condition. The time-dependent viability of the virus also plays a significant role. Here, the influence of the viability is higher at low air change rates compared with high ones, because the virus stays in the room air for a longer time period and the proportion of inactivated pathogens increase. However, the effect of time-dependent viability is not that important that a low air change rate has an overall positive effect.

To reduce the risk of infection via aerosols the necessary volume flow of virus-free air depending on the exposure time can be seen in Figure 5. This figure may be helpful to implement measures, such as increasing the virus-free air supply rate. Furthermore, the number of exposed persons must be kept in mind. An infection risk of 60% may result in one infected person in a two-person office, but in 60 infected persons in a room with 100 persons.

Predicting the infection risk via aerosols and knowing the important parameters can help in the selection of appropriate preventive actions.

## Data Availability

all data presented in the manuscript

## Acknowledgement

We thank Claudia Ruscher (State Office for Health and Social Affairs (LAGeSo), Berlin, Germany), Marius Hausner (Local Health Authority of Berlin-Mitte, Berlin, Germany), Mareike Kunze (Local Health Authority of Berlin-Mitte, Berlin, Germany), Bettina Weiss (Local Health Authority of Charlottenburg-Wilmersdorf, Berlin, Germany) and Felix Reichert (Department of infectious disease epidemiology, Robert Koch-Institute, Berlin, Germany) for providing information on outbreak B, as well as Sabine Timm (Bernau, Berlin) for providing information on outbreak K.

## Declarations

The authors received no specific funding for this work.

The authors declare no competing interests.

The authors declare that they followed the appropriate research guidelines.

